# The importance of mothers: The social transmission of COVID-19 vaccination attitudes and uptake

**DOI:** 10.1101/2024.03.06.24303875

**Authors:** Oscar Thompson, Mioara Cristea, Monica Tamariz

**Affiliations:** Independent scholar; Department of Psychology, Heriot-Watt University, Riccarton Campus, Edinburgh, UK

**Keywords:** social transmission, vertical and horizontal transmission, vaccination uptake, attitudes

## Abstract

The global fight against the COVID-19 pandemic has underscored the critical importance of widespread vaccination to mitigate the impact of the virus on public health. The current study aimed to investigate which social influences might be most important for predicting attitudes towards COVID-19 vaccination and vaccine uptake among young students in the UK. We focused on the cultural evolution and social transmission aspects, i.e., parent-to-child versus peer-to-peer, of attitudes and vaccine uptake during the COVID-19 pandemic. A sample of 192 UK students (aged 18 to 35 years old) filled in an online survey including measures for attitudes towards COVID-19 vaccination and vaccine uptake and/or intention, age, and gender. Participants were also asked about their mother’s, father’s, and best friend’s attitudes towards COVID-19 vaccination and vaccine uptake. Finally, they provided a subjective measure of the quality relationship with their parents. Overall, our results suggest that both parents and very close friends are important agents in understanding the students’ attitudes towards COVID-19 vaccination and vaccine uptake. More specifically, our findings suggest the mother’s vaccine uptake as the most salient predictor of students’ attitudes towards COVID-19 vaccination and vaccine uptake, particularly when the students disclose having a positive relationship with their parents. In cases where students’ experience negative relationship with their parents, the best friend’s vaccine uptake may supersede the mother’s influence. Despite these nuances, a general trend emerges from our data suggesting that vaccine uptake could be primarily guided by vertical transmission (i.e., parent to child). Our results have the potential to influence public health strategies, communication campaigns, and targeted interventions to enhance vaccination uptake. Identifying key social predictors can enable policymakers and health authorities to tailor vaccination promotion efforts towards mothers’ and peers’ vaccine uptake to increase overall positive attitudes and vaccine uptake among young people.

## 1 Introduction

High rates of non-vaccination pose serious risks. During COVID-19, those unvaccinated increased the strain on the NHS and may have increased the liklihood of more lethal variants emerging (1). Given looming health threats like the Avian Flu (2), understanding why some remain unvaccinated is critical. A plethora of research has explored how psychological and sociodemographic factors relate to vaccine intentions and uptake (3,4). However, the precise influence of an individual’s immediate relationships on vaccination behaviour remains under-investigated. Existing work often aggregates the association of parents and peers’ norms on vaccination intentions and uptake (5,6). By not separating the individual contribution of parents and peers, interventions are limited to a general approach focusing on changing overall norm perceptions. Delineating whether parents or peers are more salient influences for young people could focus interventions on the most important agents shaping young people’s vaccine behaviour. The present study aims to fill this gap by comparing the association of parents and peers’ norms in predicting the COVID-19 vaccination attitudes and uptake of a young UK student population. We hope to provide a more nuanced understanding of how vaccination attitudes and uptake spread within one’s social circle to inform more targeted public health interventions.

### 1.1 Social psychological models

The Theory of Planned Behaviour (TPB) (7) posits that attitudes, social norms, and perceived behavioural control predict behavioural intention, and behavioural intentions can predict behaviours (8,9). Attitudes, social norms (both injunctive and descriptive), and perceived behavioural control moderate the relationship between intention and behaviour (10,11). TPB has been used to predict various health intentions but less often fully explains behavioural change (12). Meta-analyses suggest that TPB accounts for only 19.3% to 27% of variance in health behaviours (13,14). Regarding COVID-19 vaccination, a meta-analysis indicated that attitudes (*r* = 0.48) and social norms (*r* = 0.43) are reliable predictors of intention and are hence important during vaccination (15).

However, the TPB may have limitations when used in isolation given it cannot fully explain behaviour (16). Interventions only utilising TPB factors may not be comprehensive enough to fully address behaviours, and factors outside the model need considered. For example, within the social norm’s component, the TPB assumes family and friends equally influence an individual’s intentions. In contrast, cultural transmission psychology suggests people are biased towards certain individuals within their social circle, i.e., family and friends can play different roles of importance (17,18). Understanding the dynamics between parents, peers, and behaviour could provide more targeted social interventions for improving vaccination rates.

### 1.2 Cultural evolutionary theory

Cultural evolutionary theory is concerned with the factors that shape the spread and transmission of information such as behaviours, attitudes, beliefs, values, etc in populations over time. Cultural transmission, the social transfer of information between individuals, is seldom random; in most cases, it is biased (18,19). An Individual will usually observe several attitudes and behavioural variants for the same cultural trait (e.g., different strategies to solve a problem, different opinions, or habits). The choice of which variant they will adopt is guided by several types of biases. The most relevant for this study is model-based bias (18), favouring variants that are produced by: first, people (models) who are perceived to have higher skill or success (or their proxy, social prestige) (20) and second, models who are similar to us (homophyly) (21). Model-based bias is present in adults (22–24) and children (25,26) and across cultures (27). This implies individuals may be biased to copy some social information from specific models, rather than from all models equally.

Another fundamental distinction in cultural transmission, made by Cavalli-Sforza and Feldman is between three cultural transmission modes: vertical transmission, from parents to children; oblique transmission, from other adults to younger individuals, and horizontal transmission, among similar-age relatives, friends and peers (17). Much cultural information and skills are passed on vertically, from parents to children in hunter-gatherers (28,29), in farming communities (27,30,31) as well as in industrialised societies (32). Vertical transmission predominates in early infancy and pre-school age, but parental influence then decreases in favour of peer influence, which peaks in adolescence (27,33–36).

The mode of transmission employed by a particular cultural variant has long-term cultural-evolutionary consequences. Variants that are transmitted vertically and obliquely can persist for many generations. Oblique and horizontal transmission support spread across families and populations thus accelerating the rate of cultural evolution (37,38). Variants that are transmitted exclusively horizontally will disappear once everyone in the relevant age-cohort has died out.

Moreover, the context in which we learn and transmit modulate the spread of cultural variants. We tend to transmit to peers’ information that we have learned from peers, and when we are in a position of expertise, we transmit to novices the information that we learned from experts (39). These context-congruence effects may therefore lock a behaviour in a transmission mode. Given these effects, to design effective interventions to promote desired behaviours and discourage undesirable ones it is essential that we understand the transmission modes they employ. Parental, vertical transmission may be affected by model-based biases favouring older, expert or prestigious individuals (mother and father). In contrast, horizontal transmission from friends may be biased in favour of variants learned from those like us (homophyly).

### 1.3 Social transmission

Cavalli-Sforza and Feldma (32) suggested that deep-rooted traits like political ideology and religious beliefs are more likely be socially inherited from parents than peers. However, young people may rely on peers and online sources, more than on parents for gathering health-related information (40). Looking at other health behaviours, the exercise habits of peers longitudinally predict those of young adults (41,42), whereas parents’ exercise does not (43). Health-positive behaviours may be more influenced by peers, suggesting that pro-health actions like vaccination could be determined more by peer behaviour than by parents. Regarding health-risk behaviours, alcohol consumption in adolescents (44,45) and young adults (42) is again better predicted by peer consumption than by parent consumption, at least in cross-sectional studies (46). Smoking in early adolescents is also best predicted by peers’ smoking behaviour (47). As such, social context and age matters. Young students are surrounded by new influences, different from their parents, which may render peer behaviour a more important model during COVID-19 vaccination.

Longitudinally, however, parents predict maladaptive health behaviour such as alcohol consumption (48,49), smoking (49) and resistance to wearing seatbelts (42) better than peers. Despite peer influence increasing in adolescence, parent-child conflicts may diminish into young adulthood (50), making parental influence salient again, later in life. Maladaptive health behaviours of young people could, in the long run, be better predicted by parents. Vaccine attitudes and uptake in young people might follow a similar maladaptive behavioural pattern, being more influenced over time by parents than peers.

Both parents’ and peers’ behaviours are therefore important predictors of a young student’s health behaviours. Parental behaviour may continue to be a significant predictor for young adults, even when exposed to new social landscapes such as a university setting (42). However, the importance of either influence can vary across different behaviours and contexts. Consequently, COVID-19 vaccination uptake among students could equally be biased more by either parents or peers’ behaviour and attitudes. To alter COVID-19 vaccination attitudes and increase uptake among students, it is crucial to discern the most important social influences biasing vaccine hesitancy.

### 1.4 COVID-19 Vaccination

Much COVID-19 vaccination research has focused on the extent to which attitudes regarding the vaccine’s safety, efficacy, and perceived importance are consistent predictors of vaccination intention and uptake (5,51). With this importance of attitudes, understanding how parents and peers may individually predict a student’s attitudes is imperative for trying to indirectly influence their vaccination uptake.

Research on factors predicting vaccine intentions and uptake either combines family and friends as a single normative influence or deals with only family or only friends. Specifically, among students, vaccine intentions (52) and uptake (53) have been associated with a perception of peers’ willingness to receive the vaccine. Similarly, having an unvaccinated family member or friend has been associated with an increased likelihood of the student being unvaccinated (54).

Importantly, however, the influence of parents versus peer norms has not been directly compared, leaving it unclear whether one might be more impactful than the other. This is especially relevant for interventions using peers as vaccine models, e.g., (2). If parental association prevails, such peer-focused interventions could be undermined, preferring to bias their vaccine attitudes and uptake through observing their parents.

In one study that compared parents and peers’ influence on young adolescents, the authors found that parent and peer COVID-19 vaccination attitudes and intentions both predicted US adolescent vaccination intentions, but parents norms explained twice the variance of intention over peer norms (56). These effects, however, could be different in an older sample of students, who are more exposed by peer influence, and parental influence could decline (27,35). The role of parents on COVID-19 vaccination in a young student population, at an age and in a context where peer influence is at a peak, has not been examined. It’s therefore crucial to investigate the comparative association of parents and peers within this population.

Within parents, fathers and mothers may indeed play different roles. During the COVID-19 lockdowns in the UK, lower maternal compliance with guidelines was a stronger predictor than paternal behaviour of reduced compliance among young adults, suggesting a predominant influence of mothers during COVID-19 (57).Thus, separating mothers’ and fathers’ behaviour will permit interventions to focus on the most salient parental influence. Furthermore, the association was moderated by relationship quality, with increased family arguments weakening the mother-child compliance association (57). Parental behaviour was also not significantly related to young adults who were living away from home during lockdowns. As such, parents could be less important for students, whom may live with peers, with an effect of both one’s household and parental relationship quality in determining the social influences on attitudes and uptake of COVID-19 vaccination.

### 1.5 Research questions and objectives

This study aims to address the gap in the literature identified above, focusing on the cultural evolution and social transmission aspects of attitudes towards and uptake of COVID-19 vaccines during the COVID-19 pandemic. We aim to identify which social agent—a mother, father, or best-friend—is the strongest predictor of students’ vaccination attitudes and uptake. Directional hypotheses were not made given the scant literature of parents and peers during COVID-19 vaccination. Instead, we explored the data to suggest where relationships might emerge.

An individual’s COVID-19 vaccination attitudes are consistent predictors of vaccination intentions and uptake (6,51). Therefore, we first investigated who the strongest predictors of student’s attitudes were to understand how to change young people’s COVID-19 beliefs via more indirect social modelling interventions. Next, we predicted student vaccine uptake (i.e., whether the student is vaccinated or not) to explore who could be the largest risk factor for young people rejecting a COVID-19 vaccine. We included relationship quality of students and their parents as covariance to investigate if it moderated parents’ association with the student. Lastly, using a heritability analysis, we explored how COVID-19 vaccine attitudes and uptake are transmitted across one’s whole circle.

## 2 Methods

### 2.1 Participants

We conducted a UK-wide survey on vaccination attitudes and uptake. A total of 346 participants accessed the survey, of which 192 met our criteria (18-35 years of age, UK students). Gender distributions showed a higher proportion of females than males (females = 126, males = 57, other = 7), with a Mean age = 21.0 years. Convenience sampling was used to gather participants. Advertisements were placed on Reddit, Facebook, Twitter, LinkedIn, and Instagram - inviting participants to join a study requesting UK students to give their opinion and behaviours surrounding on COVID-19 vaccination.

In the UK, COVID-19 vaccines were first offered to residents and staff of older adult care homes In December 2020. It wasn’t until June 2021 that it was offered to 21- and 22-year-olds, and 18 July to 18–20-year-olds. Booster doses were offered to all adults 6 months after the first dose, meaning by January 2022 all adults 18 or over had been offered a second or booster dose. Participant recruitment took place between 19^th^ July 2021 and 27^th^ January 2023. Recruitment advertisements were placed online and through social media at two-time windows: (1) June-August 2021 and (2) November 2022-January 2023. Participants were able to access links between the two windows, but most responses were received during the two advertisement periods.

### 2.2 Instruments

Participants were asked how many vaccines they had received (0, 1 or 2 or more). Vaccination intention for unvaccinated participants was measured using one item on a 10-point Likert scale, i.e., “Now that a coronavirus vaccination is available, how likely is it that you will have one?” (0 - ‘extremely unlikely’ and 10 -‘extremely likely’). Attitudes towards COVID-19 vaccination were measured using four bi-polar adjectives (on 10-point Likert scales, i.e. 0-very ineffective to 10-very effective; 0 - very unsafe to 10-very safe; 0 - very unimportant to 10 - very important, and 0-very negative to 10-very positive). Similarly, they stated how many vaccines they thought each of the agents had (their mother, father and best friend), and finally, they were asked to rate their perceptions of each agent’s attitude towards COVID-19 vaccination on the same bipolar adjective scales (e.g., “What is your impression on what the following people THINK about a COVID-19 vaccination…?”). Agents were presented in randomised order for each participant. Attitude components were all highly correlated, and so a composite COVID-19 vaccination attitude variable was made with high reliability for the student (α = .95), mother (α = .97), father (α = .97), friend (α = .95). Finally, participants provided a subjective measure of their parents’ relationship quality, using a measure “how positive is your relationship with your mother/father?” with 1 being very negative and 5 being very positive. We asked some additional questions that were not used in analysis. The full survey is available in the Supplementary Information.

### 2.3 Procedure

An advertisement online presented participants with a link that took them to a Qualtrics survey. In the survey, they first were asked to provide informed consent to participate. Participants who consented proceeded to complete the survey described above. Finally, they were fully debriefed.

### 2.4 Ethics statement

The study was granted ethical approval by the School of Social Sciences Ethics Committee, Heriot-Watt University (2021-1268-4512). Participant consent was obtained electronically at the start of the survey. Participants read a sheet informing them of the project aims, the task and their rights, and were asked to select “I agree” or “I do not agree”. Only those who agreed were directed to the rest of the survey described above.

### 2.5 Analytical Procedure

R was used to conduct all statistical analyses. Data was imported from excel and cleaned and analysed through R. The data and code is available at https://github.com/oscarthompsoncodes/TCT-Vaccine-2024. Details of the exact analytical procedures for handling the data is found in the results section “Testing Linearity Assumptions” as procedure was decided upon after testing various data assumptions. Our sample size was N =192, but full data was not available for all variables for each participant. As such, some analyses had less participants used in the model due to data incompleteness, and the number of observations used in each analysis is stated throughout the tables.

## 3 Results

All the data were analysed together, given that separate analyses of the early and later datasets showed similar distributions of vaccination intentions, uptake, and attitudes.

### 3.1 Descriptive statistics and correlations

Table 1 presents descriptive statistics. Most respondents were vaccinated and had positive attitudes towards the COVID-19 vaccination. Vaccine intention was very low, indicating most of those unvaccinated had no intention of being vaccinated. Students’ relationships with their mother and father were also positive overall.

**Table 1.**
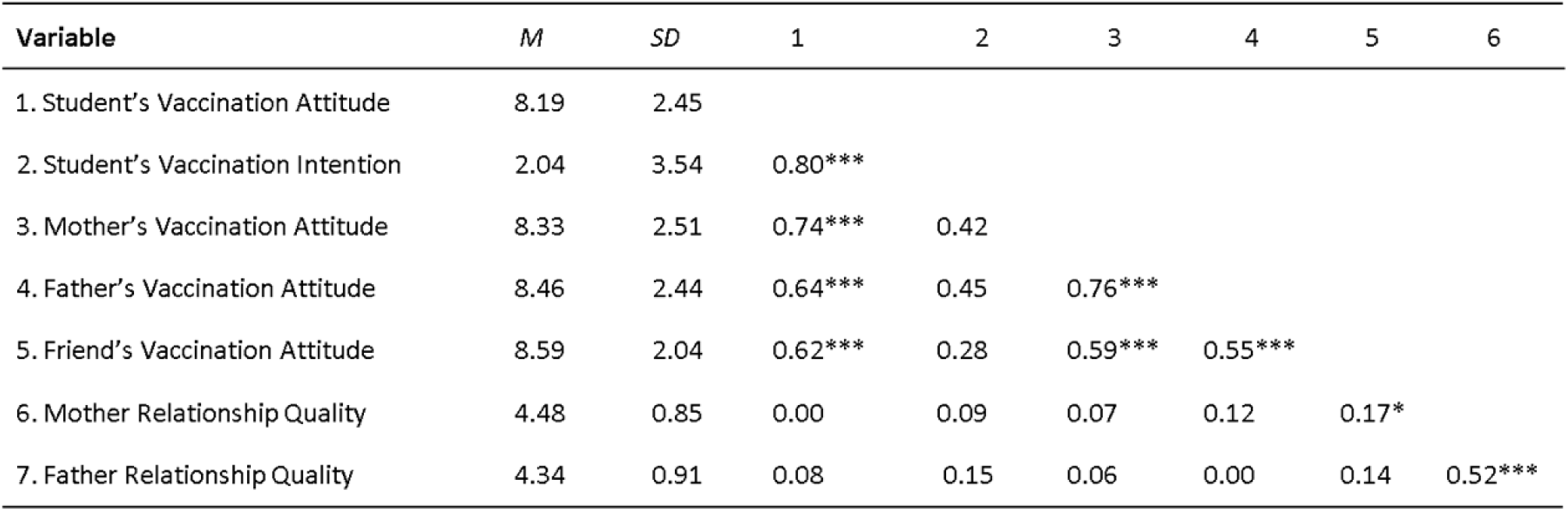
Means, standard deviations, and correlations between students’ attitudes towards COVID-19 vaccination and intentions and each agents attitudes towards COVID-19 vaccination. *Note.* * *M* and *SD* are used to represent mean and standard deviation, respectively. Spearman correlations were used given variables were non-normally distributed. Student COVID-19 Vaccine Intention refers to the intention to receive a COVID-19 vaccination for those who were unvaccinated. Mother and Father Relationship Quality refers to how positive the student perceived their relationship with their mother and father. ** Indicates *p* < .05. ** indicates *p* < .01. *** indicates p < .001.

| Variable | *M* | *SD* | 1 | 2 | 3 | 4 | 5 | 6 |
| --- | --- | --- | --- | --- | --- | --- | --- | --- |
| 1. Student's Vaccination Attitude | 8.19 | 2.45 |  |  |  |  |  |  |
| 2. Student's Vaccination Intention | 2.04 | 3.54 | 0.80*** |  |  |  |  |  |
| 3. Mother's Vaccination Attitude | 8.33 | 2.51 | 0.74*** | 0.42 |  |  |  |  |
| 4. Father's Vaccination Attitude | 8.46 | 2.44 | 0.64*** | 0.45 | 0.76*** |  |  |  |
| 5. Friend's Vaccination Attitude | 8.59 | 2.04 | 0.62*** | 0.28 | 0.59*** | 0.55*** |  |  |
| 6. Mother Relationship Quality | 4.48 | 0.85 | 0.00 | 0.09 | 0.07 | 0.12 | 0.17* |  |
| 7. Father Relationship Quality | 4.34 | 0.91 | 0.08 | 0.15 | 0.06 | 0.00 | 0.14 | 0.52*** |

*Note.*
\*\* Indicates $p < .05$ . \*\* indicates $p < .01$ . \*\*\* indicates $p < .001$ .

Of the 27 respondents who were not vaccinated; 21 had no or little intention of getting a vaccine (vaccine intention scores 0, 1 or 2), 1 was neutral (vaccine intention score of 5); and 4 had a strong intention of getting a vaccine (vaccine intention scores 9 or 10).

Table 3 includes the correlations between each agent’s vaccination uptake. Here the highest correlation was observed was between the mother and the student, followed by the mother and the father.

To complement our correlation analysis, we calculated social heritability between the agents. In biology, a trait (such as height, skin colour, intelligence) is heritable when offspring resemble their parent more than another random individual from the population regarding that trait. In culture, a socially learned trait such as a behaviour or attitude is heritable if an agent (e.g., students) resemble a cultural model (e.g., their mothers) more than random individuals for the relevant behaviour or attitude.

We conducted Monte-Carlo analyses to quantify the social heritability of vaccination attitude and behaviour scores as the z-score of the veridical correlation between two agents (e.g. students and their mothers) in a distribution of 10,000 scrambled correlations (e.g., each student paired with a random mother from the sample). Heritability results (Fig 1), despite being dependent on the composition of the population, nevertheless contribute evidence of transmission between the agents as the causal process supporting similarity. However, our analysis is agnostic regarding the precise mechanisms for transmission, which could be biological (genetic or epigenetic inheritance), social (learning, imitation, teaching, and uni- or bi-directional, e.g. from father to student, but also from student to father), or a combination of both.

**Figure 1.**
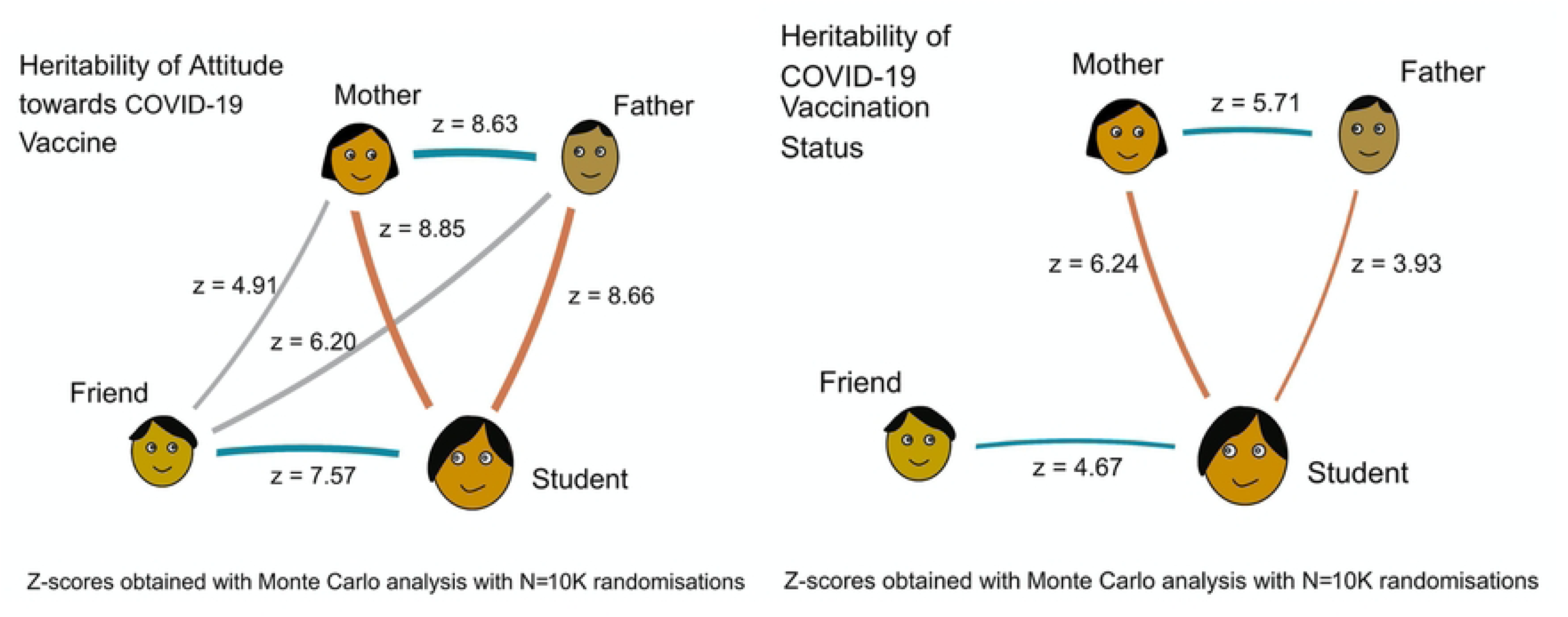
Heritability of attitudes towards COVID-19 vaccination (left) and COVID-19 vaccination uptake (right) for all pairs of agents in our sample. Only z-scores > 3.1, corresponding to p < 0.001 are shown.

Results suggest that the population is clustered into homogeneous groups including family and friends who share the same COVID-19 vaccination attitudes, but when it comes to vaccination uptake, clusters are not as homogeneous, but students nevertheless resemble their friend’s, their father’s, and particularly their mother’s behaviour.

### 3.2 Regressions

#### 3.2.1 Testing linearity assumptions

To ensure robust analysis, we used box plots, QQ plots, and residual plots for outlier detection. Although potential outliers were initially identified, these corresponded having a lower number of participants with very negative vaccination attitudes. We determined the dataset did not contain any meaningful outliers. We therefore decided to retain the complete dataset for subsequent regression analyses.

Normality plots and QQ plots revealed non-normal data distribution, attributed to COVID-19 vaccination attitudes being polarized as very negative or very positive. The data failed to meet the assumption for linear models, and log transformations did not normalise the data. COVID-19 vaccination attitudes were highly collinear (see Table 2). Dormann and colleagues (58) advocates a correlation coefficient threshold of 0.7 for predictor variables to preserve accurate coefficient estimation. Given attitudes approached this threshold, results were interpreted cautiously for attitudes. GLMs were preferred for handling collinearity and nonlinearity (58).

**Table 2.**
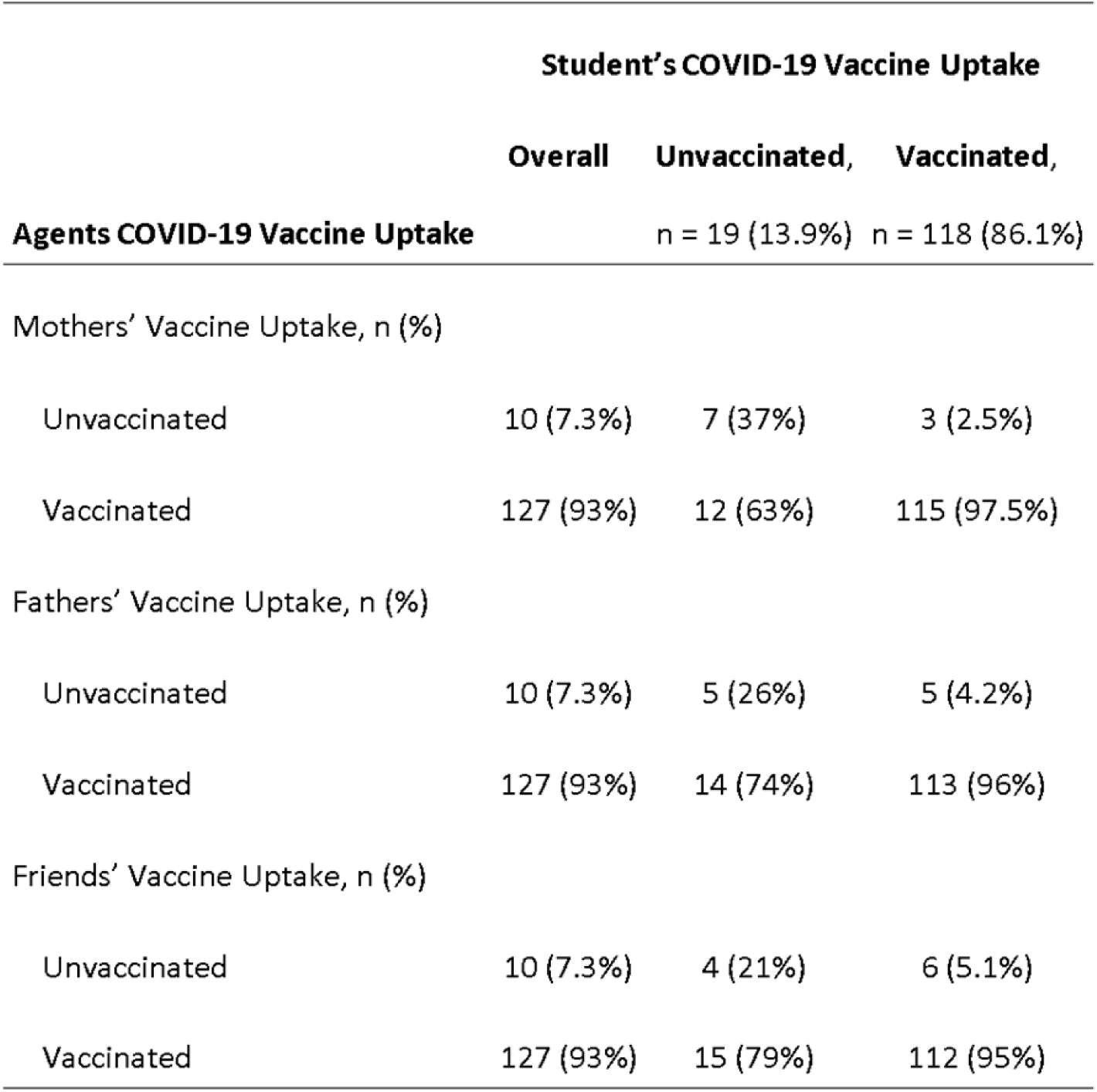
COVID-19 Vaccine Uptake of the student and agents and their similarities (n = 137). Only complete cases are included, so participants who did not provide data for the vaccination uptake of their mother, father and friend were removed.

|  | Student's COVID-19 Vaccine Uptake |  |  |
| --- | --- | --- | --- |
|  | Overall | Unvaccinated,<br>n = 19 (13.9%) | Vaccinated,<br>n = 118 (86.1%) |
| **Agents COVID-19 Vaccine Uptake** |  |  |  |
| Mothers' Vaccine Uptake, n (%) |  |  |  |
| Unvaccinated | 10 (7.3%) | 7 (37%) | 3 (2.5%) |
| Vaccinated | 127 (93%) | 12 (63%) | 115 (97.5%) |
| Fathers' Vaccine Uptake, n (%) |  |  |  |
| Unvaccinated | 10 (7.3%) | 5 (26%) | 5 (4.2%) |
| Vaccinated | 127 (93%) | 14 (74%) | 113 (96%) |
| Friends' Vaccine Uptake, n (%) |  |  |  |
| Unvaccinated | 10 (7.3%) | 4 (21%) | 6 (5.1%) |
| Vaccinated | 127 (93%) | 15 (79%) | 112 (95%) |

Vaccine uptake data was unaffected by self-report bias and only have low to moderate correlations between each agent (see Table 3). For these reasons, uptake data may be more informative than attitude data, alongside using the quality of relationship between students and parents as covariates, which are largely unrelated to all agents’ attitudes (see Table 1).

**Table 3.** Correlations between agents’ vaccination uptake. * Indicates p < .05. ** indicates p < .01.

| Variable | 1 | 2 | 3 |
| --- | --- | --- | --- |
| 1. Student Vaccine Uptake |  |  |  |
| 2. Mother Vaccine uptake | .48** |  |  |
| 3. Father Vaccine uptake | .33** | .48** |  |
| 4. Friend Vaccine Uptake | .36** | .10 | .25** |

Furthermore, a p-value was not pre-determined nor adjusted for multiple comparisons. Rather the study aims to explore the data fully and point to where meaningful relationships could exist. Consequently, the significance, regression coefficient and powers of model predictors will be considered in interpretation.

#### 3.2.2 Predictors for Students’ Attitudes towards Vaccination

To investigate predictors of students’ COVID-19 vaccination attitudes, three GLM models with Poisson links were executed, as outlined in Table 6. Model 1 assessed the influence of other agents’ attitudes; model 2 examined the association with other agents’ vaccine uptake; and model 3 combined both. Analyses were restricted to complete data sets containing information for students and their three specified agents (mother, father, friend).

GLM with Poisson link was used in these models by treating the attitude variables as count data. As Poisson link requires a positive skew, student and others’ attitudes were transformed to a positive skew. Each variable was taken away from 11 to reverse the distribution. Lower attitude values therefore indicate a positive vaccination attitude, and vice versa.

In model 1 (Table 4), the students’ attitude towards COVID-19 vaccination were modelled by each agent’s attitude towards COVID-19 vaccination. A quasi-poisson model was conducted to check for dispersion. At 0.6, the data was not over-dispersed. The father and the best-friend attitudes were the only significant predictors, and the father’s attitudes was the strongest predictor. A 10% change in fathers’ attitudes was associated with an 11.2% change in students’ attitudes, while a 10% change in the friends’ attitudes was associated with a 10.6% change. The difference between the fathers’ and friends’ attitudes is thus negligible in meaningful predictive strength.

**Table 4.** Student’s attitudes towards COVID-19 vaccination predicted by the agents’ attitude towards COVID-19 vaccination and their vaccine uptake. Each column represents a separate Poisson regression where the outcome variable was the student’s attitudes towards COVID-19 vaccination. CI indicate 95% CIs. R^2^ Nagelkerke was the goodness of fit test used for the Poisson regressions. The coefficients used are incidence rate ratios because a Poisson GLM was used. These were converted from their log form for easier interpretation.

| *Predictors* | Model 1:<br>Others' Attitudes |  |  | Model 2:<br>Others' Uptake |  |  | Model 3:<br>Attitudes & Uptake Combined |  |  |
| --- | --- | --- | --- | --- | --- | --- | --- | --- | --- |
|  | *IRR* | *CI* | *p* | *IRR* | *CI* | *p* | *IRR* | *CI* | *p* |
| Intercept | 1.38 | 1.14 – 1.67 | **0.001** | 11.39 | 7.34 – 16.88 | **<0.001** | 0.76 | 0.39 – 1.52 | 0.440 |
| Mother's Attitude | 1.05 | 0.98 – 1.12 | 0.169 |  |  |  | 1.06 | 0.95 – 1.18 | 0.255 |
| Father's Attitude | 1.12 | 1.04 – 1.20 | **0.002** |  |  |  | 1.15 | 1.04 – 1.28 | **0.010** |
| Friend's Attitude | 1.06 | 1.00 – 1.13 | **0.039** |  |  |  | 1.06 | 0.99 – 1.13 | 0.086 |
| Mother's Uptake |  |  |  | 0.43 | 0.28 – 0.69 | **<0.001** | 1.41 | 0.54 – 3.55 | 0.473 |
| Father's Uptake |  |  |  | 1.00 | 0.60 – 1.72 | 0.994 | 1.67 | 0.64 – 4.70 | 0.312 |
| Friend's Uptake |  |  |  | 0.50 | 0.35 – 0.75 | **<0.001** | 0.71 | 0.42 – 1.20 | 0.191 |
| Observations | 106 |  |  | 128 |  |  | 105 |  |  |
| $R^2$ Nagelkerke | 0.828 | | | 0.450 | | | 0.860 | | |

Fig 2 shows a high clustering of data points in the bottom left, indicative of the larger amount of very positive attitudes towards COVID-19 vaccination. Moreover, the regression lines are very similar. The association of each agent’s attitudes with the student’s attitudes are very similar, suggesting that attitudinal variables are interrelated and collectively contribute to the predictive power of the model. Consequently, it becomes challenging to discern the individual association of each agent’s attitude.

**Figure 2.**
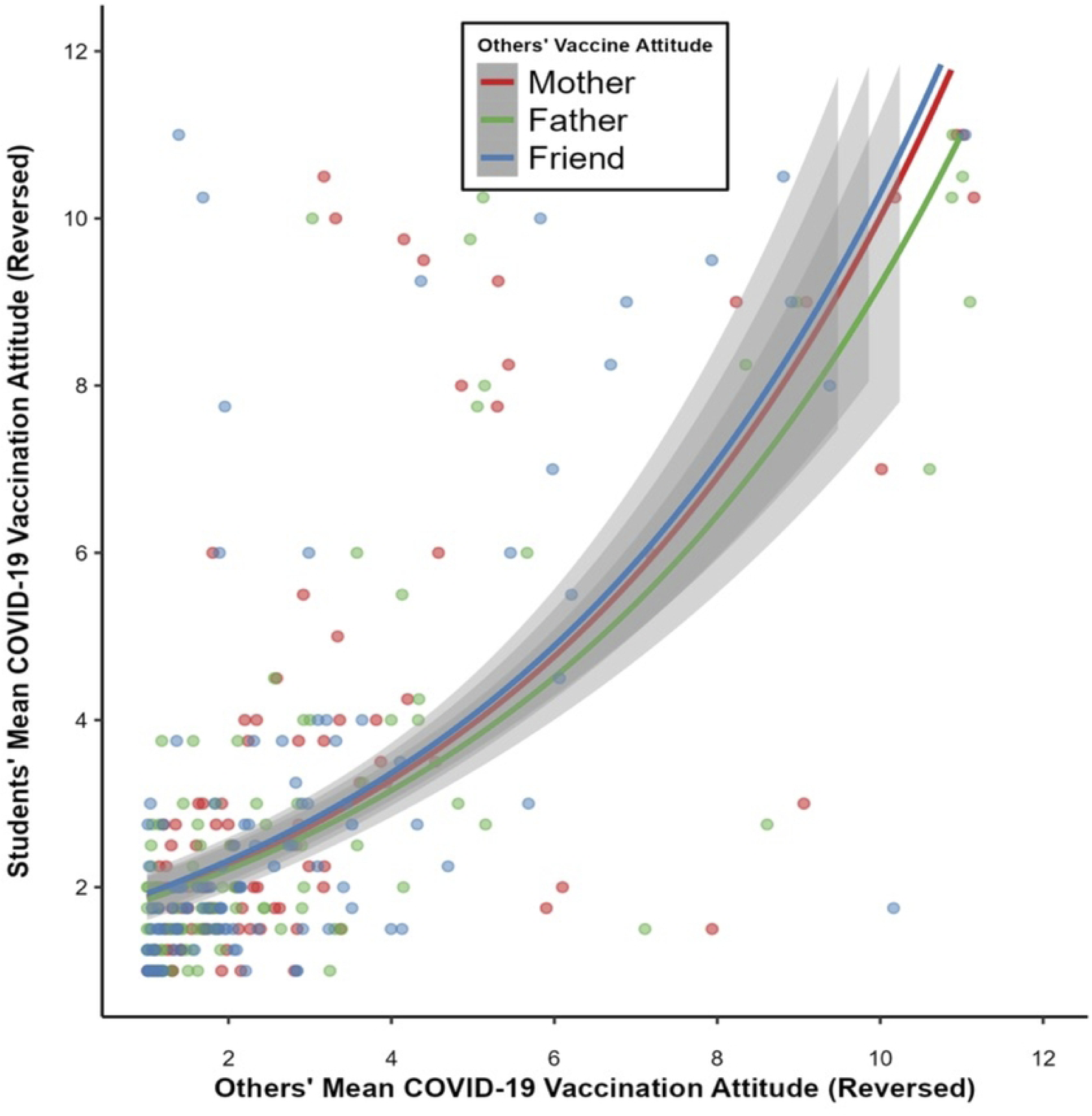
Plot of the Poisson regression of the student’s COVID-19 vaccination attitudes modelled by other’s COVID-19 vaccination attitudes. Each line indicates a separate Poisson regression run, with each agent’s attitudes towards COVID-19 vaccination predicting the student’s attitudes towards COVID-19 vaccination. Attitude values reversed: 1 = very positive attitude towards COVID-19 vaccination, and 10 = very negative attitude towards COVID-19 vaccination.

In model 2 (Table 4) students’ attitudes towards COVID-19 vaccination were modelled by other agents’ COVID-19 vaccine uptake. The model was over-dispersed (dispersion = 1.62, with a dispersion criterion of <= 1). The Quasi-Poisson model was used to account for this overdispersion.

The mother’s vaccine uptake was the strongest predictor for student’s attitude towards COVID-19 vaccination, followed by the friend’s uptake. The father’s uptake was non-significant. The mother’s vaccine uptake predicted student attitudes with an IRR = 0.43. This means having a vaccinated mother was associated with the student having 57% more positive attitudes compared to students who had an unvaccinated mother. Conversely, having a vaccinated friend was associated with a student having 50% more positive vaccination attitudes, compared to students with an unvaccinated friend. The mothers’ vaccine uptake therefore accounts for 7% more variance in predicting student attitudes than the friends’ vaccine uptake.

Fig 3 illustrates these findings. The student’s attitudes are universally positive towards vaccination when the mother (M = 8.40, CI = 8.05-8.75), father (M = 8.32, CI = 7.92-8.72), and friend (M = 8.51, CI = 8.16-8.85) are vaccinated. Conversely, the student has the lowest vaccination attitudes when their mother is unvaccinated (M = 3.88, CI = 0.77-6.98), followed by their friend (M = 4.40, CI = 2.21-6.58), and attitudes are highest when the father is unvaccinated (M = 5.19, CI = 1.82-8.55).

**Figure 3.**
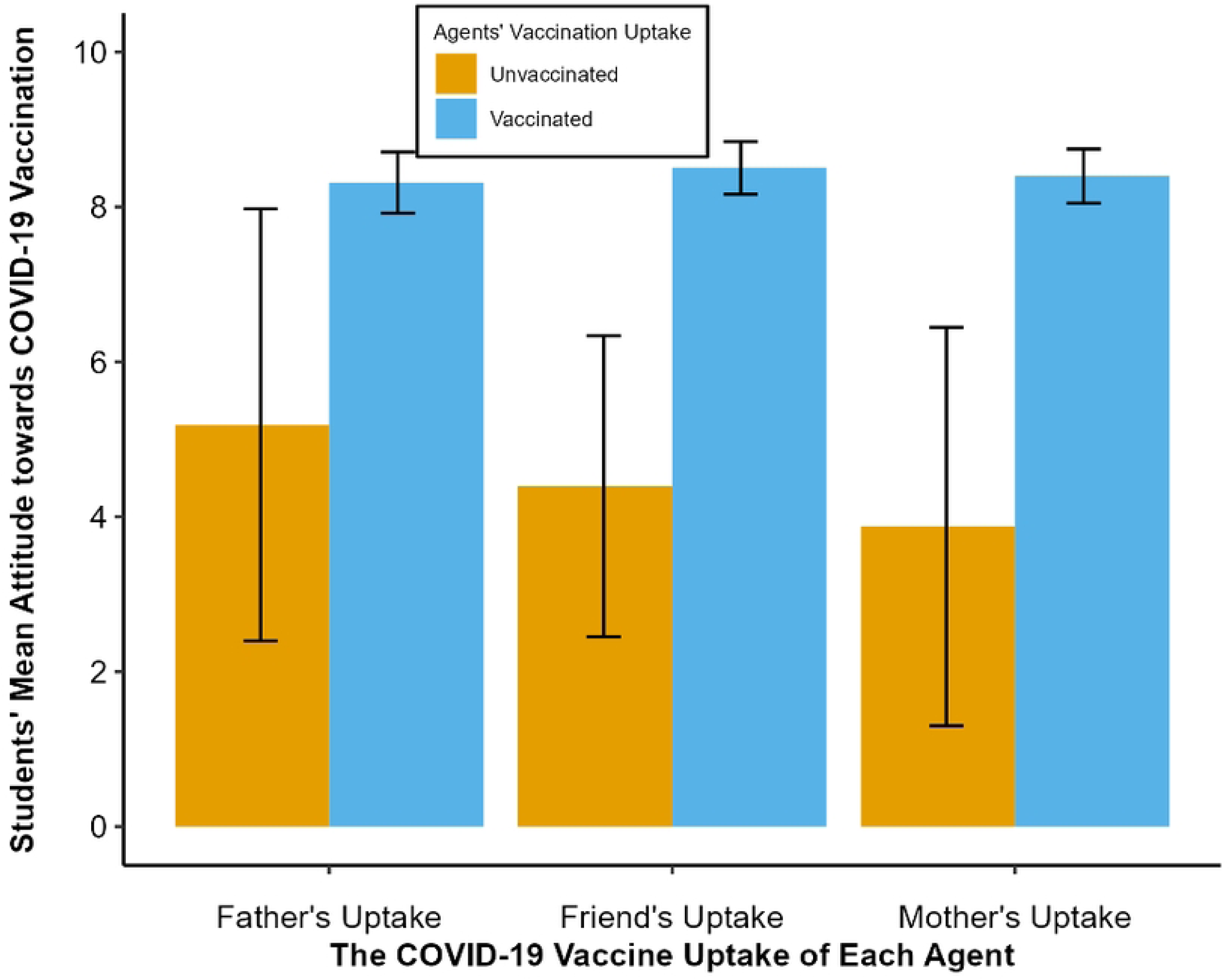
Means and 95% CI of the student’s COVID-19 vaccination attitudes when each person is either vaccinated or unvaccinated. Vaccination uptake refers to the COVID-19 vaccination uptake of others. Attitude towards COVID-19 vaccination variable scaled went from: 1 = a very negative attitude towards vaccination to 10 = a very positive attitude towards vaccination.

The large confidence intervals suggest that the precise estimates of the mother or friends’ coefficients may be unreliable. But still they suggest the direction that the mother’s uptake is a stronger predictor than the father’s or friend’s uptake: having an unvaccinated mother was associated with the lowest vaccination attitudes, followed by having an unvaccinated friend.

Model 3 summarises how others’ attitudes and uptake together predict student attitudes. Note, the goodness of fit is highest in this model. Only the father’s attitudes towards COVID-19 vaccination were a significant predictor of student attitudes. None of the agents’ vaccine uptake was significant predictors of student attitudes. However, with attitudes being highly collinear, it is unlikely the fathers’ attitude accounted for much unique variance, and this finding is interpreted more cautiously.

#### 3.2.3 Predictors of students’ vaccination uptake

It may be of greater interest to understand the predictors of vaccine uptake. Utilising the same predictors as in Table 4, three new regression models were constructed to explore how others’ attitudes and uptake best predicted student vaccine uptake. The results are outlined in Table 5.

**Table 5.** Student’s uptake of COVID-19 vaccination predicted by the COVID-19 vaccination attitudes and uptake of parents and their friend. Each model here was a binomial logistic regression. R^2^ Tjur was the goodness of fit measure used for the logistic regression models. The model originally produced log OR but we transformed these to OR for easier interpretation. Note that some confidence intervals were very high (e.g., model 6, Mothers’ Uptake.) However, this was not due to modelling or formatting issues, but was the real values obtained from the models. With highly collinear variables, it’s expected there would be some unusual CIs for GLM models, especially given the combined model was more underpowered with a lower observation to variable ratio. Given we are exploring the data, we left these values in as is. We follow this procedure for the full article.

| *Predictors* | **Model 4:**<br>**Others' and Student's Attitudes** |  |  | **Model 5:**<br>**Others' Uptake** |  |  | **Model 6:**<br>**Attitudes and Uptake Combined** |  |  |
| --- | --- | --- | --- | --- | --- | --- | --- | --- | --- |
|  | *Odds Ratios* | *CI* | *p* | *Odds Ratios* | *CI* | *p* | *Odds Ratios* | *CI* | *p* |
| Intercept | 0.52 | 0.04 – 6.89 | 0.606 | 0.09 | 0.01 – 0.76 | **0.037** | 0.03 | 0.00 – 1.72 | 0.181 |
| Mother's Attitude | 0.74 | 0.33 – 1.27 | 0.382 |  |  |  | 0.36 | 0.08 – 0.99 | 0.105 |
| Father's Attitude | 1.47 | 0.84 – 3.03 | 0.239 |  |  |  | 1.44 | 0.67 – 3.64 | 0.386 |
| Friend's Attitude | 0.94 | 0.60 – 1.35 | 0.739 |  |  |  | 0.79 | 0.33 – 1.46 | 0.513 |
| Student's Attitude | 1.48 | 0.97 – 2.31 | 0.060 |  |  |  | 2.52 | 1.34 – 6.00 | **0.013** |
| Mother's Uptake |  |  |  | 16.64 | 3.34 – 99.83 | **0.001** | 3087.93 | 1.05 – 2.33e8 | 0.083 |
| Father's Uptake |  |  |  | 1.90 | 0.25 – 11.23 | 0.499 | 5.99 | 0.01 – 505.12 | 0.472 |
| Friend's Uptake |  |  |  | 3.88 | 0.65 – 18.78 | 0.105 | 0.04 | 0.00 – 5.88 | 0.245 |
| Observations | 106 |  |  | 137 |  |  | 105 |  |  |
| R<sup>2</sup> Tjur | 0.319 |  |  | 0.238 |  |  | 0.459 |  |  |

Vaccine uptake was not significantly predicted by the student’s own or other’s attitudes towards COVID-19 vaccination (model 4, Table 5). Fig 4 shows the students and others’ attitudes when the student was either vaccinated, or unvaccinated. When the student was vaccinated, their mothers’ (M = 8.85, CI = 8.55 - 9.16), fathers’ (M = 8.90, CI = 8.57 - 9.23), friends’ (M = 8.90, CI = 8.57 - 9.23, SD = 1.77), and own attitudes (M = 8.72, CI = 8.45 - 8.99) were all similarly high. When the student was unvaccinated, the student’s attitude seemed lowest (M = 3.57, CI = 1.89 - 5.26), which would be expected given attitudes would be in correspondence to their own behaviour. But the mother (M = 5.18, CI = 3.41 - 6.94), father (M = 5.79, CI = 3.92 - 7.66), and friends’ (M = 7.08, CI = 5.61 - 8.55) attitudes did not seem to differ substantially, perhaps explaining why there was no significant predictors. When a student is unvaccinated, no agents’ attitudes are uniquely predictive of student uptake.

**Figure 4.**
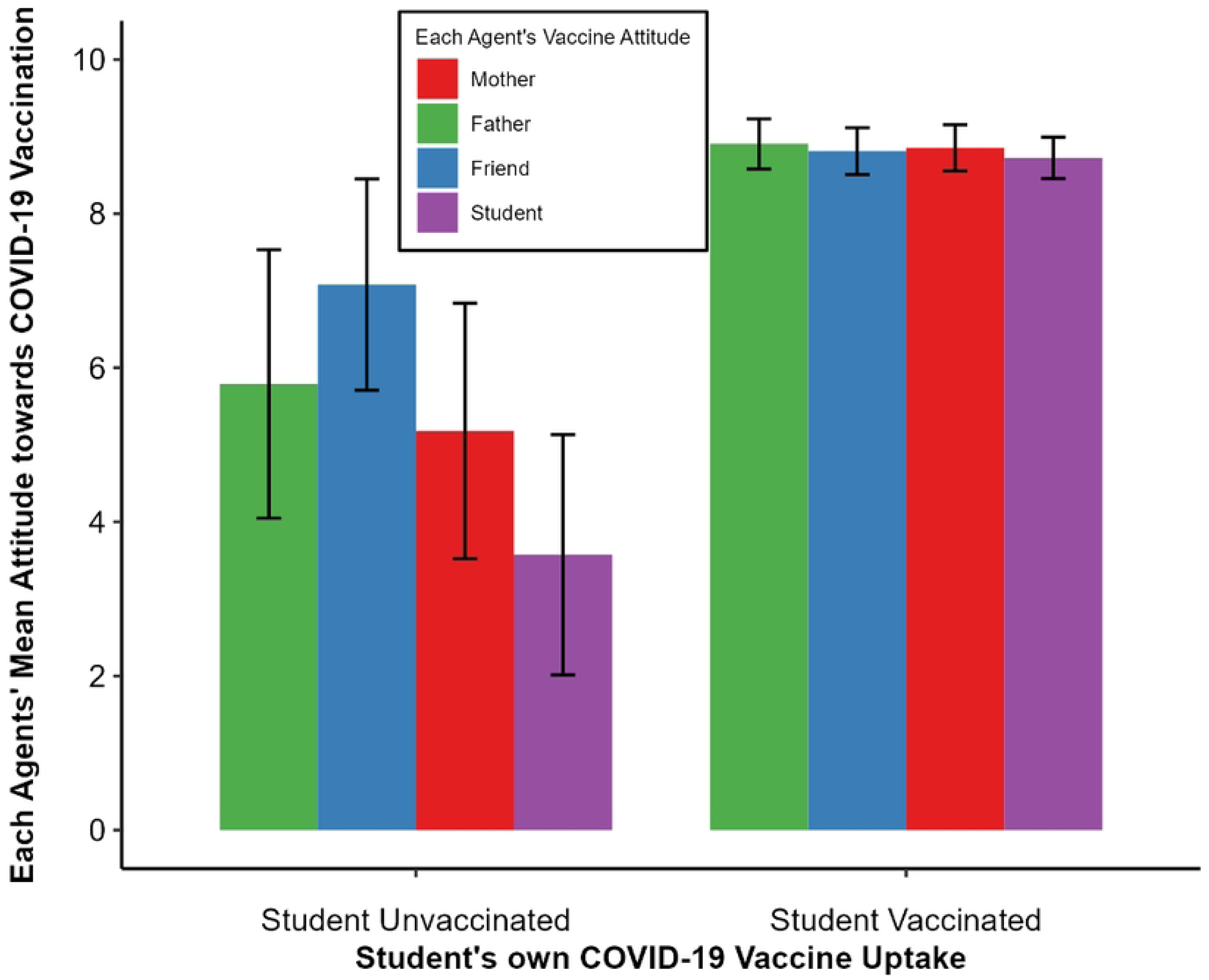
Mean and 95% CIs of the COVID-19 vaccination attitude values of others and the student when the student is either vaccinated or unvaccinated. Attitude towards COVID-19 vaccination variable scaled went from: 1 = a very negative attitude towards vaccination to 10 = a very positive attitude towards vaccination

As was the case with predicting students’ attitudes towards COVID-19 vaccination, investigating the relationships between other’s uptake and student’s uptake may be more informative for understanding how vaccination uptake is predicted, because it does not rely on self-reports. Model 5 in Table 5 summarises the findings. The mother’s vaccine status was the only significant predictor of whether a student was vaccinated. Having an unvaccinated mother was associated with a 16 times increased likelihood of the student being unvaccinated themselves. However, the large confidence intervals indicate the precision of this OR is uncertain.

Table 2 explores these findings. Vaccinated students’ vaccine uptake was equally like all the other agents’ vaccine uptake, ranging from 95-97%. The results were different for unvaccinated students. Here, 37% of the time the mother was also unvaccinated, versus 26% for father, 21% for friend, supporting the potential importance of mother’s uptake for student uptake, especially when the mother is unvaccinated.

Combined model 6 included both the attitudes and uptake of other agents in predicting students’ vaccine uptake. The log binomial model indicated the student’s attitude was the sole predictor. While this model better controls for the covariance across variables, this model was again less powered, with only 108 observations across 8 variables. We suggest this model is thus not powered enough to dismiss the importance of the mothers’ uptake observed in model 5.

#### 3.2.4 Relationship quality as a covariate

Aksoy found the transmission of lockdown compliance norms during the pandemic was higher from mothers to their children in families that argued less (54). Thus, the strength of association from parents to their children might depend on relationship quality. We re-ran the models including the positivity of mother and father relationships as covariates (see Methods section).

##### 3.2.4.1 Predicting student’s attitudes

We first explored how others’ attitudes and uptake predicted a students’ COVID-19 vaccination attitudes when controlling for the quality of the relationship between students and their parents’ as covariance. Table 6 outlines the outcomes of these models. The same structure of models was used as in Tables 4 and 5.

Model 7 (Table 6) summarises the findings. The model’s dispersion was in range D = 0.66. When considering the quality of the parents’ relationships, the friend’s attitude was the only significant predictor of student attitudes.

In model 8 (Table 6) we next examined how vaccine uptake of the student was predicted by the agents’ vaccine uptake when the quality of the relationships with parents was a covariate. An unvaccinated friend now emerged as the strongest predictor of student attitudes, followed by an interaction effect between the mother’s vaccination uptake and relationship quality (Fig 5). Students with vaccinated mothers exhibited positive vaccination attitudes irrespective of relationship quality. In contrast, students with unvaccinated mothers showed diverging attitudes based on the quality of the relationship; positive relationships led to lower vaccination attitudes, whereas negative relationships led to higher attitudes. A student’s attitude towards vaccination is more likely to align with the vaccine uptake of their mother only if they get on well with her. Student’s attitudes are not universally predicted by maternal vaccination uptake. As such, friends’ vaccination uptake may be the better predictor of student’s vaccine attitudes when controlling for student-parent relationship quality.

**Figure 5.**
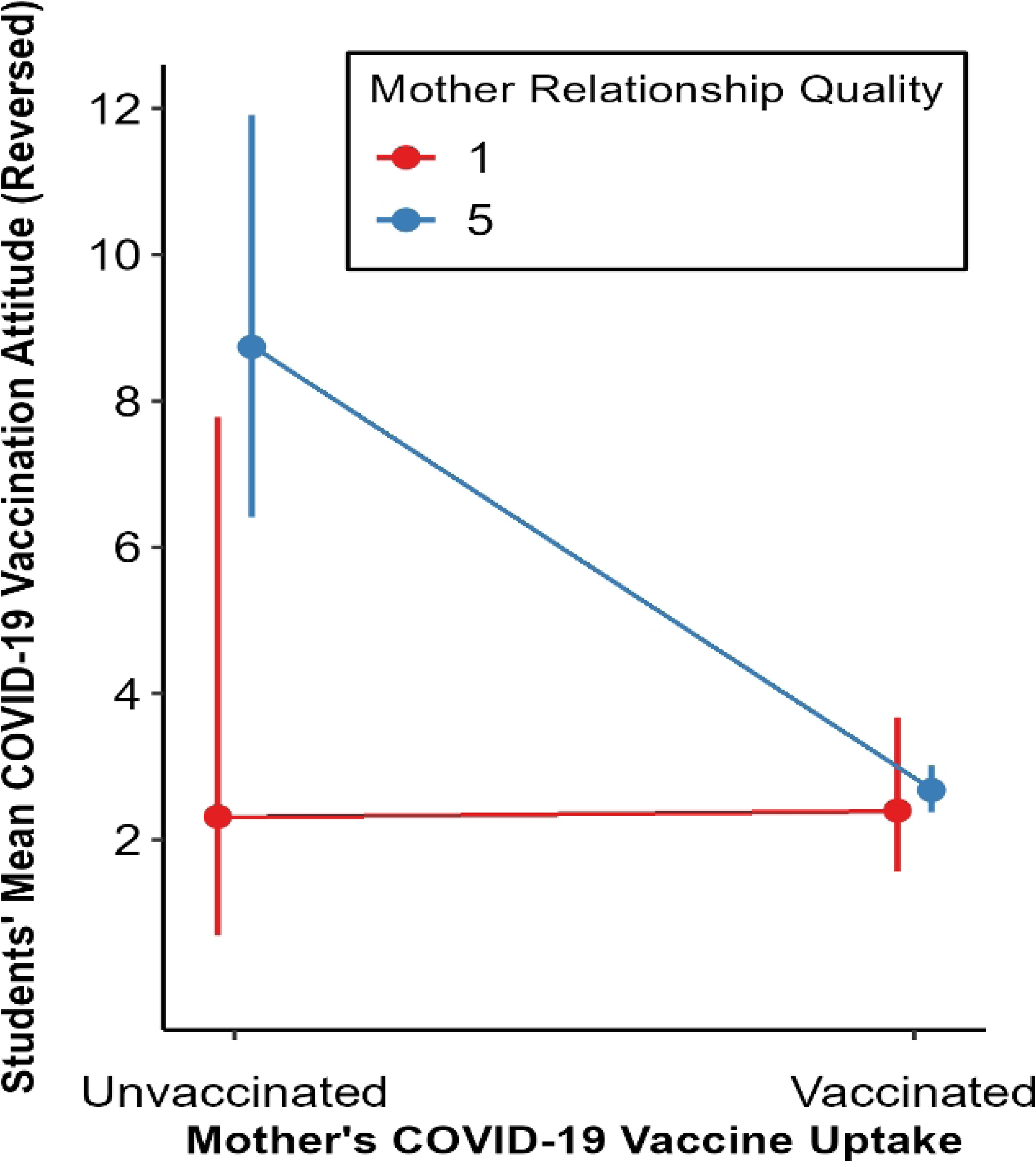
Student attitudes for those with a vaccinated and unvaccinated mother, with positivity of the mother’s relationship as an interaction effect. Error bars show 95% CIs. Students’ attitude values reversed: 1 = very positive attitude towards COVID-19 vaccination, and 10 = very negative attitude towards COVID-19 vaccination. Mother relationship quality was continuous, but here split as an interaction term, using the minimum (1 – very negative relationship) and maximum (5 – very positive relationship) as factor levels for a simpler interpretation.

In the combined model 9 (Table 6), when the quality of the father’s relationship was included as a covariate, the father’s attitude was no longer significant, and the vaccine uptake of the friend was the sole predictor. Thus, the father’s attitude emerging as a significant predictor may have depended on the student having a positive relationship with them. This might suggest that friends’ attitudes and uptake are more informative than the father’s attitudes. Thus, having an unvaccinated friend may be the most important predictor of student’s vaccination attitudes.

**Table 6.** Student’s attitudes towards COVID-19 vaccination predicted by the agents’ COVID-19 vaccination attitudes and uptake, including parental relationship quality as covariance. Rel Qual refers to relationship quality. In the original combined model with all agents’ attitudes and behaviours for predicting students’ attitudes (model 3, Table 4), only the father’s attitudes were a significant predictor. Hence, relationship quality was included as a covariate only for the mother’s and father’s attitudes, not their vaccination uptake.

| *Predictors* | **Model 7:**<br>**Others' Attitudes** |  |  | **Model 8:**<br>**Others' Uptake** |  |  | **Model 9:**<br>**Attitudes and Uptake Combined** |  |  |
| --- | --- | --- | --- | --- | --- | --- | --- | --- | --- |
|  | *Incidence*<br>*Rate Ratios* | *CI* | *p* | *Incidence Rate*<br>*Ratios* | *CI* | *p* | *Incidence Rate*<br>*Ratios* | *CI* | *p* |
| Intercept | 2.07 | 0.72 – 5.75 | 0.169 | 0.66 | 0.02 – 12.43 | 0.792 | 4.96 | 1.27 – 18.91 | **0.020** |
| Mother's Attitude | 1.14 | 0.70 – 1.85 | 0.598 |  |  |  | 0.85 | 0.52 – 1.38 | 0.522 |
| Father's Attitude | 0.92 | 0.60 – 1.42 | 0.691 |  |  |  | 0.88 | 0.59 – 1.32 | 0.533 |
| Friend's Attitude | 1.07 | 1.01 – 1.14 | **0.015** |  |  |  | 0.99 | 0.92 – 1.06 | 0.794 |
| Mother's Attitude x Rel Qual | 0.99 | 0.90 – 1.09 | 0.851 |  |  |  | 1.06 | 0.96 – 1.18 | 0.259 |
| Father's Attitude x Rel Qual | 1.04 | 0.95 – 1.13 | 0.395 |  |  |  | 1.06 | 0.97 – 1.15 | 0.202 |
| Mother's Rel Qual | 1.00 | 0.75 – 1.34 | 0.982 | 1.42 | 1.01 – 2.14 | 0.061 | 0.86 | 0.65 – 1.13 | 0.272 |
| Father's Rel Qual | 0.91 | 0.71 – 1.18 | 0.470 | 1.37 | 0.78 – 2.66 | 0.312 | 0.85 | 0.68 – 1.07 | 0.154 |
| Mother's Uptake |  |  |  | 2.84 | 0.52 – 20.22 | 0.257 | 1.66 | 0.76 – 3.52 | 0.195 |
| Father's Uptake |  |  |  | 2.27 | 0.14 – 60.08 | 0.591 | 1.53 | 0.71 – 3.55 | 0.302 |
| Friend's Uptake |  |  |  | 0.48 | 0.35 – 0.67 | **<0.001** | 0.45 | 0.28 – 0.73 | **0.001** |
| Mother's Uptake x Rel Qual |  |  |  | 0.64 | 0.42 – 0.93 | **0.028** |  |  |  |
| Father's Uptake x Rel Qual |  |  |  | 0.84 | 0.42 – 1.51 | 0.579 |  |  |  |
| Observations | 99 |  |  | 120 |  |  | 98 |  |  |
| R<sup>2</sup> Nagelkerke | 0.822 |  |  | 0.437 |  |  | 0.867 |  |  |

##### 3.2.4.2. Students’ vaccine uptake

Secondly, we modelled how others’ attitudes and vaccine uptake predicted a students’ COVID-19 vaccination uptake when including parental relationship quality as a covariate (Table 7).

**Table 7.** Student attitudes modelled by the attitudes and behaviours of others, including parental relationship quality as covariance. We did not re-run a combined model including relationship quality as a covariate because in model 6 (Table 5), the combined model predicting vaccine uptake, neither parents’ attitudes or uptake were significant predictors.

| *Predictors* | **Model 10:**<br>**Others' Attitudes** |  |  | **Model 11:**<br>**Others' Uptake** |  |  |
| --- | --- | --- | --- | --- | --- | --- |
|  | *Odds Ratios* | *CI* | *p* | *Odds Ratios* | *CI* | *p* |
| Intercept | 18.34 | 0.00 – 1.34e11 | 0.794 | 3351.63 | 0.00 – 3.19e14 | 0.476 |
| Mother's Attitude | 2.43 | 0.12 – 98.35 | 0.593 |  |  |  |
| Father's Attitude | 0.48 | 0.02 – 6.32 | 0.597 |  |  |  |
| Friend's Attitude | 0.89 | 0.51 – 1.36 | 0.635 |  |  |  |
| Mother's Attitude x Rel Qual | 0.80 | 0.36 – 1.51 | 0.528 |  |  |  |
| Father's Attitude x Rel Qual | 1.32 | 0.79 – 2.46 | 0.333 |  |  |  |
| Mother's Rel Qual | 6.85 | 0.03 – 5795.47 | 0.533 | 0.24 | 0.01 – 1.51 | 0.172 |
| Father's Rel Qual | 0.08 | 0.00 – 7.40 | 0.338 | 0.34 | 0.00 – 6.74 | 0.608 |
| Mother's Vaccine Uptake |  |  |  | 0.02 | 0.00 – 171.89 | 0.428 |
| Father's Vaccine Uptake |  |  |  | 1.87 | 0.00 – 1.62e7 | 0.953 |
| Friend's Vaccine Uptake |  |  |  | 3.70 | 0.50 – 21.05 | 0.155 |
| Mother's Uptake x Rel Qual |  |  |  | 5.47 | 0.65 – 118.35 | 0.151 |
| Father's Uptake x Rel Qual |  |  |  | 0.99 | 0.03 – 121.23 | 0.996 |
| Observations | 102 |  |  | 128 |  |  |
| R<sup>2</sup> Tjur | 0.274 |  |  | 0.276 |  |  |

In model 10 (Table 7), when predicting student vaccine attitudes, the attitudes of agents were non-significant (without the covariate, the father’s attitudes were the sole significant predictor). Similarly, in model 11 (Table 7), none of the agents’ uptake variables were significant predictors of student uptake (whereas the mother’s uptake was without the covariate). Therefore, when controlling for the quality of parent-student relationship, not one agent accounts for unique vaccine uptake.

### 3.3 Gender interactions

As the sample was mostly female (N = 66.3% females) maternal importance may have existed because females displayed a bias towards the same-gender parents. Female adolescents copied their mother’s COVID-19 compliance behaviours more than male adolescents (57). Each model was re-run with gender as a covariate. The pattern of the findings did not change across any of the models. As such the greater percentage of females in the sample did not bias the findings. Importantly, the salience of the mother does not depend on student gender.

## 4 Discussion

As COVID-19 vaccination has rolled out, young people have remained some of the most hesitant age-group within the UK (59). Much research has suggested how family’s and friends’ COVID-19 vaccination norms are important in predicting COVID-19 vaccination intention and uptake. However, social interventions may be made more effective by targeting the specific social connection – namely, parents or peers – which is more important in shaping the young person’s vaccine uptake. Therefore, this paper’s objective was to examine how three agents’ – mothers, fathers and best friends--attitudes and uptake predict students’ COVID-19 vaccination attitudes and uptake. These findings could be used to infer which agent might have the greatest influence on young people’s vaccine uptake to inform interventions.

### 4.1 Predicting student’s attitudes towards COVID-19 vaccination

Our first model predicted students’ COVID-19 vaccination attitudes by others’ COVID-19 vaccination attitudes. The mother’s, father’s and friend’s COVID-19 vaccine attitudes predict the student attitudes to the same degree (Fig 2). However, we observe that all agents’ attitudes, including the student’s, were highly collinear. This collinearity means that the significant coefficient of the father and friend’s attitudes on student’ attitudes are not to be overemphasised with no one agent being more important than the others.

This collinearity may be partly due to mothers, fathers and friends’ attitudes being reported by the students, who may be biased to perceiving those who are most close to them as having the same evaluations as each other regarding vaccine importance, safety, effectiveness, and positivity. Moreover, they may be biased to believe these three agents have similar attitudes to themselves (60). Besides these biases, collinearity of attitudes among all four agents (student, their mothers, fathers, and friends) indicates that our population may be polarised into homogeneous groups. Here, students surrounded by parents with positive attitudes would have access to or choose to befriend or marry people with positive attitudes, and the same for negative attitudes.

Our second model included other agents’ vaccine uptake, a more objective measure, as predictors. Having an unvaccinated mother was associated with the most negative COVID-19 vaccination attitudes, followed by having an unvaccinated friend. The mother’s vaccine uptake accounted for 7% more variance in student attitudes than the friend’s vaccine uptake. The father’s vaccine uptake was not significant. When developing intervention campaigns to improve student’s attitudes towards vaccination, the focus should therefore be prioritising the vaccination of mothers as well as friends, but less so fathers.

Our third combined model including the COVID-19 vaccination attitudes and uptake of the mother, father, and friend showed only the father’s attitude predicted the student’s attitudes. However, given findings from the previous models, issues with collinearity and the lower power of the larger combined model, this result should not be overstated. Overall, these models suggest that a student’s attitude towards vaccine is most influenced by their whole close social circle with the mothers and friends’ vaccination uptake being most important.

Most COVID-19 vaccine research has not separated out the individual contribution of parents and peers (5,15)Instead, participants are asked about their perception of both their family and friends’ attitudes and intentions towards COVID-19 vaccination as a single overall variable. This overall perception has found to be consistently important for predicting people’s vaccine intentions and attitudes, e.g., (5,51). Yet, our separation of mother versus father versus friend nuances these results to underline specific biases towards individual agents, namely mothers.

### 4.2 Predicting student’s COVID-19 vaccine uptake

Our fourth model indicated that individual attitudes, including those of the students themselves, were not unique predictors of students’ COVID-19 vaccine uptake. In our fifth model, including only other agents’ vaccine uptake, the mother’s uptake was the only significant predictor of student uptake. An unvaccinated mother may therefore be the most important social predictor of a young person’s failure to get vaccinated.

Our sixth, combined model, however, found the student’s own attitudes as the sole predictor of their vaccine uptake, challenging the mother’s importance. This is in line with some studies (6), but not with others (56), which indicate that our close social agents can have a stronger association in predicting vaccine intention than our own attitudes.

Once again, the lower power of the combined model and the collinearity issue suggests the importance of mothers’ uptake on student’s uptake should not be discounted. Combined with the importance of the mothers’ non-vaccination in predicting negative vaccination attitudes, maternal influence remains as a potentially crucial risk factor for vaccine attitudes and uptake.

This is in line with Kecojevic and colleagues’ findings (54) who showed that students who reported having an unvaccinated family member or friend were more likely to be unvaccinated. Other studies focusing on peers’ norms suggest peers may influence young people’s attitudes and uptake of COVID-19 vaccination (52,53). Our findings suggest while peer norms may be important, interventions relying solely on peers may be insufficient, as maternal influence could prevail over peer norms in shaping the student’s vaccine uptake.

Our results also align with those of Rogers and colleagues (56) who showed that parent norms accounted for twice the variance in adolescents’ COVID-19 vaccine intention than peer norms. Their sample was younger than ours (Mean age = 14.69 years). At this age parents are likely the main deciders regarding vaccination. Our findings, in a slightly older sample (Mean age = 21.0 years) suggest that parents may continue to shape COVID-19 vaccination uptake into young adulthood despite being surrounded by peer influence in a student context. Note, we did not record the living circumstances of students, yet findings have previously suggested young adults’ association with parental COVID-19 guideline compliance increases when living at home (54). As such our findings cannot determine whether the association was due to living around parents versus peers.

Additionally, the perceived influence of peers may be confounded by students merely selecting friends with similar vaccination uptake. Given our young sample, and the age-based rollout of the vaccination in the UK, participants were likely offered the vaccine after they had already observed their parents reject or accept the vaccine. There was therefore a greater opportunity to observe parental vaccine decisions and imitate their behaviour compared to peers.

We extended Rogers and colleagues’ findings (56) by showing that, among parents, the mother’s uptake was a singularly important predictor of student uptake. Similarly, Aksoy (57) showed a mother’s lockdown compliance behaviour predicted UK young adults’ compliance behaviour, whereas a father’s behaviour was less or non-significant. The importance of mothers might be expected, given children spend more time with their mother growing up in social activities and educational events (61),have more communication with their children than fathers (62). Thus, the mother may be of central importance during unfamilar situations like pandemics, both acting as models for young people through health regulation compliance and making important vaccination decisions.

### 4.3 Social transmission research

Health risk behaviours are sometimes more likely to be predicted by parental behaviour than peer behaviour in young adults, e.g., alcohol consumption (48,63), or smoking (49), although these results are not always supported, e.g. for alcohol consumption (42). In opposition, peer behaviour is a better predictor for exercise habits (41–43) and health searching (64)Thus, research must continue to investigate the effects of parents versus peers and indeed of mothers versus fathers, to understand which social agents shape behaviour and attitudes.

### 4.4 Cultural evolutionary Psychology

An explanation for the importance of mother’s vaccine uptake on student’s uptake may be that important, deep-rooted traits like politics and religion tend to be transmitted vertically, from parents to children, rather than horizontally, among peers (65). COVID-19 vaccination hesitancy is related to right-wing political views (66,67) and increased religiosity (68). Amid the uncertainty of vaccination decisions, young people may have looked towards modelling the behaviour of their parents, for whom they’ve previously followed similar fundamental ideologies. Consequently, vertical transmission could be the more important transmission modality for COVID-19 vaccination.

Moreover, vertically transmitted behaviours and attitudes –unlike horizontally transmitted ones– can persist for many generations (39). This means that the influence of unvaccinated mothers may be transmitted down the generations. Therefore, addressing vaccine hesitancy especially among mothers may be crucial to stop the transmission of anti-vaccination attitudes and behaviours.

### 4.5 Heritability analyses

Heritability analysis generally aligns with the regression results. In addition, they offer a view of the distribution of attitudes and behaviours across our sample. For attitudes, all agents are very similar to each other, indicating the existence of homogeneous groups within which all agents have similar attitudes towards the COVID-19 vaccine. This is reflected in the fact that friends are very similar to parents, although they are not directly related. Friends are less like parents in terms of vaccine uptake, suggesting uptake is not as polarised as attitudes. Therefore, interventions should still consider one’s whole circle for most effectively shaping vaccine uptake, and not singularly focus on one social agent.

### 4.6 Relationship as Covariance

When controlling for parental relationship quality (how positive or negative the student reported their mother and father relationships were), the friend’s predictive power increased. When predicting student vaccination attitudes, an unvaccinated friend was associated with the most negative attitudes. Having an unvaccinated mother was only associated with more negative student vaccination attitudes for those who had a positive relationship with their mother. Furthermore, the friend’s uptake was the sole predictor for student attitudes in the combined model. But no agent’s attitude or uptake were significant predictors of student uptake when controlling for relationship quality.

The importance of parents’ behaviour may therefore be dependent on how well the child and parents get on. In COVID-19, a mother’s compliance with lockdown guidelines predicted their young adult children’s compliance only if the children lived with them and they argued less (57). Our sample reported very positive relationships with their mother (M = 4.48/5) and father (4.34/5), suggesting the importance of parents could be overestimated in our results. This implies that findings by Rogers and colleagues (56), who suggested parent norms were more important than peer norms for vaccination, may have been different if they had included relationship quality in their design. Their adolescent participants may have been biased in favour of parents simply because they had a positive relationship with them. Consequently, research must consider how well a young person gets on with other social agents to most accurately determine why a social agent might appear to be of greatest importance.

### 4.7 Practical applications of findings

If our results are verified by further studies, interventions aimed at improving vaccine uptake could target the agents who are found to be most important. For instance, leveraging vaccination of mothers to act as behavioural models could be a more specific approach for changing attitudes and increasing uptake in young adults. In situations where children do not get along with their mothers, friends could play a more important role in influencing uptake. Therefore, parent-based interventions and friend-based interventions complement each other, and neither should be ruled out.

### 4.8 Limitations

There are limitations to the current study that need to be acknowledged. Firstly, we used a correlational design, which does not allow to infer causality. We obtained merely measures of similarity, so we cannot say whether an agent directly influenced the uptake and attitudes of the student. For instance, it is possible that similarity between mother and student is caused by the student influencing the mother. For the friend, inferring directionality is even less plausible, since friendship can involve both people influencing each other equally.

This study’s aim was to establish patterns of social transmission of information in a small social network. However, we relied on similarity metrics between different agents. Similarity of attitudes and behaviours can be caused by factors other than transmission including sharing a social environment; for parents and children, sharing genes; and, for friends, assortative mating whereby we select friends who are like us. Future studies can tease out these confounds to understand the unique contribution of social transmission in attitudes and behaviours towards vaccination.

We relied on student’s perception of their parents’ and friend’s COVID-19 vaccination attitudes and uptake. Even if they do not accurately reflect the real attitudes and behaviours, perceptions may be what leads to someone changing their own attitudes and behaviours. Furthermore, within the complete vaccine uptake data we had, (N =137) we had only 27 unvaccinated participants (19.7% unvaccinated). Although unbalanced, this proportion is representative of the rate of non-vaccination in the UK.

## 5. Conclusion

This study aimed to compare the predictive power of parents and peers on COVID-19 vaccination attitudes and uptake among students to determine who could be the greatest source of influence for vaccination uptake. Our results support the view that both one’s close family and friends are important. Moreover, findings point to the mother’s vaccine uptake as the most salient predictor of student’s uptake and attitudes, particularly when the students get on well with their parents. In cases of poorer parent-child relationships, friend’s vaccine uptake may supersede the mother’s influence. Despite these nuances, a general trend emerges suggesting that vaccine uptake could be primarily guided by vertical transmission (i.e., parent to child). In vaccination, the influence of young people’s mothers and fathers is understudied yet may be potentially more important than studying solely peer influence. Therefore, interventions should be finely tuned to utilising students’ mothers to increase vaccine uptake, whereas peers could be more important for those with poorer maternal relationships. Future research confirming the importance of mothers could be pivotal for optimising strategies to mitigate the continual vaccine hesitancy across pandemics in this vulnerable demographic.

## Data Availability

All data files are available from the github database. Access here: https://github.com/oscarthompsoncodes/TCT-Vaccine-2024

## Acknowledgements

OT thanks the Carnegie Trust for funding through a Sumer Internship.

